# Legacy immunity from prior smallpox vaccination and serological evidence of asymptomatic mpox transmission in a West African population

**DOI:** 10.1101/2025.07.08.25330960

**Authors:** Adam Abdullahi, Ifeanyi Omah, Reshma Kassanjee, Sophia Osawe, Martin Edun, Fehintola Ige, Haruna Wisso, Edyth Parker, Onikepe Folarin, Aniekwe M. Isaac, Ummasalma Aliyu Saulawa, Lourdes Ceron-Gutierrez, Anise Happi, Olga Sokolova, Rosemary Audu, Sani H. Aliyu, Christian Happi, Babatunde Lawal Salako, Rainer Doffinger, Alash’le Abimiku, Ravindra K. Gupta

**Affiliations:** International Research Centre of Excellence, Institute of Human Virology, Abuja, Nigeria; Cambridge Institute of Therapeutic Immunology & Infectious Disease (CITIID), Department of Medicine, University of Cambridge, Cambridge, UK; Takemi Program in International Health, Harvard T.H Chan School of Public Health, Boston, MA, USA; Institute of Ecology and Evolution, University of Edinburgh, The King’s Buildings, Edinburgh EH9 3FL, UK; Department of Parasitology and Entomology, Nnamdi Azikiwe University, Awka, Nigeria; Centre for Integrated Data and Epidemiological Research, School of Public Health, University of Cape Town, Observatory, South Africa; Nigerian Institute of Medical Research, Yaba, Lagos, Nigeria; Insitute for Genomics and Global Health, Redeemer’s University, Ede, Osun, Nigeria; Faculty of Natural and Applied Sciences, Umaru Musa Yar’adua University, Katsina, Nigeria; Addenbrooke’s Hospital, Cambridge University Hospitals NHS Foundation Trust, Cambridge Biomedical Campus, Cambridge, UK; Institute of Human Virology, University of Maryland School of Medicine, Baltimore, USA; College of Medicine, University of Ibadan, Ibadan, Nigeria; Africa Health Research Institute, Durban, South Africa; Hong Kong Jockey Club Global Health Institute, Hong Kong

## Abstract

The 2022 multi-country mpox outbreak, driven by MPXV Clade IIb has spread globally to over 100 countries, posing a new threat to global public health. Smallpox vaccination was discontinued in 1980 following global elimination with recent reports demonstrating evidence of asymptomatic infections particularly in individuals unlikely to have received a smallpox vaccine. To determine whether residual vaccination-derived immunity still shapes infection risk and to characterise population level burden of covert and asymptomatic transmission in a West African population, we combined serological analysis with phylogenetic analysis. We first quantified IgG binding to six MPXV antigens (A29L, A35R, B6R, D6L, H3L and M1R) in 176 Nigerian adults comprising of 75 healthcare workers sampled in 2021 and 101 community volunteers sampled in 2023 using a six-plex Luminex assay. Using a stringent ≥4-antigen reactivity threshold, at baseline, 24/176 (13.6%) participants met definition of MPXV seropositivity likely due to previous vaccination. Most participants with MPXV seropositivity were born before 1980, suggestive of residual smallpox immunity. Magnitude-breadth analysis scores were 2-fold higher in pre-1980 cohort relative to post-1980 cohort. In a subset of individuals (n=153) with follow-up samples over a median of 9 months, 5/153 (3%) showed evidence of silent asymptomatic exposure determined by antibody boosting and characterised by ≥2-fold increases in both total magnitude and breadth scores antibodies against ≥ 4/6 tested antigens all of which were MPXV seronegative individuals at baseline and mostly males. Participant level kinetics showed B6-specific titres boosted most strongly (median 11-folds), followed by M1R (6.2-folds) and A35R (5.2-folds) highlighting their potential utility as sero-surveillance markers. Complementary phylogenetic reconstruction of 105 Nigerian MPXV genomes showed two epidemic phases with over-dispersion (k≈0.3), indicating transmission sustained by sporadic superspreading despite high degree of dead-end infections. Serological and genomic evidence points to sustained asymptomatic transmission of MPXV in healthy individuals in the African setting, thereby informing public health interventions.

## Introduction

Mpox is a zoonotic disease caused by the mpox virus (MPXV), a viral member of the Poxviridae family from the Orthopoxvirus genus.^1,2^ Historically endemic to Central and West Africa, the recent global resurgence of MPXV in endemic and non-endemic regions resulted in its declaration as a Public Health Emergency of International Concern (PHEIC) in both 2022 and 2024.^3^ Unlike previous outbreaks, the 2022 outbreak, was driven by the MPXV Clade IIb, an evolutionarily distinct, pathogenically attenuated lineage with enhanced human to human transmissibility^4–9^. Thus, it has been associated with mild and asymptomatic disease which makes detection difficult and enhances silent transmission.^10–21^

A critical factor in shaping susceptibility to MPXV is residual immunity derived from historic smallpox vaccination, the only immunisation campaign in history to have eradicated a human disease and was discontinued globally in 1980 following eradication.^22^ MPXV and variola virus (agent of smallpox) share multiple conserved structural and immunogenic viral proteins such as A29L, B6R, and M1R, implicated in viral entry and immune recognition^23–32^. Individuals who were born before 1980 likely received a smallpox vaccine during the WHO-led Smallpox Eradication Programme (1966–1980)^33–36^, although global coverage was heterogenous with estimated range ≈7-60%^37^. These campaigns utilised various live-attenuated strains of vaccinia virus (VACV) stains, including the *Lister strain* (widely used across Africa, Europe, and Asia) and the *EM-63 strain* (used in the Soviet Union and parts of Africa).^38–40^ These vaccines induced broad, cross-protective immunity against Orthopoxviruses, including MPXV.^23,26,27^

While historical smallpox vaccination conferred cross-protective immunity against Orthopoxviruses,^41–50^ its global cessation has resulted in a growing global cohort of immunologically naïve individuals lacking immunity. Waning immunity in the post-smallpox era has coincided with the emergence of new MPXV transmission patterns characterised by recurrent outbreaks especially in endemic low-resource settings of sub-Saharan Africa where immunological surveillance remains limited. There is evidence of subclinical, covert and asymptomatic MPXV infection across high-resource settings including reports from USA, France, Japan and Belgium particularly in high-risk populations ^3,10–19,51^. However, whether the immunity gap has shaped transmission dynamics over time in a West African population remains poorly understood due to limited data.

To address this, we conducted two complementary analyses. First, we collected serum samples from two independent cohorts: healthcare workers in Lagos, Nigeria from 2021^52^ and the general population in Abuja, Nigeria in 2023^53^ to evaluate MPXV-specific antibody responses. In parallel, we deployed genomic methods using publicly available whole virus genome sequences to infer epidemiological dynamics. Our data firstly reveal signatures of residual smallpox immunity among individuals likely vaccinated against smallpox; secondly, we observe serological and genomic evidence suggestive of undetected community transmission of MPXV during the SARS-CoV-2 pandemic.

## Results

### Study population and cross-sectional population-level MPXV seropositivity

Our study population comprised two independent cohorts recruited during the COVID-19 pandemic to assess SARS-CoV-2 vaccine responses in Nigeria (**Figure 1**). The population comprised a total of n=176 participants (timepoint 0; T0) stratified as follows: **cohort 1**: Healthcare workers (HCW, n=75; enrolled during the SARS-CoV-2 vaccination campaign in Lagos in March 2021) and **cohort 2**: general population (n=101; enrolled during community-level vaccination campaign in Abuja in Jan 2023). The median age of participants was 37 years (IQR: 29-43) and gender was balanced (**Table 1**). A subset of these participants (n=153) had longitudinal samples available for testing (cohort 1: n= 75; and cohort 2: n=78; **T1**; median of 9 months after T0). This dual-cohort design allows us to capture the potential impact of occupational exposure to MPXV and age-related differences to MPXV-specific antibody responses, particularly in the context of historical smallpox vaccination in Nigeria which was the known origin of the 2022 outbreak.

**Figure 1.**
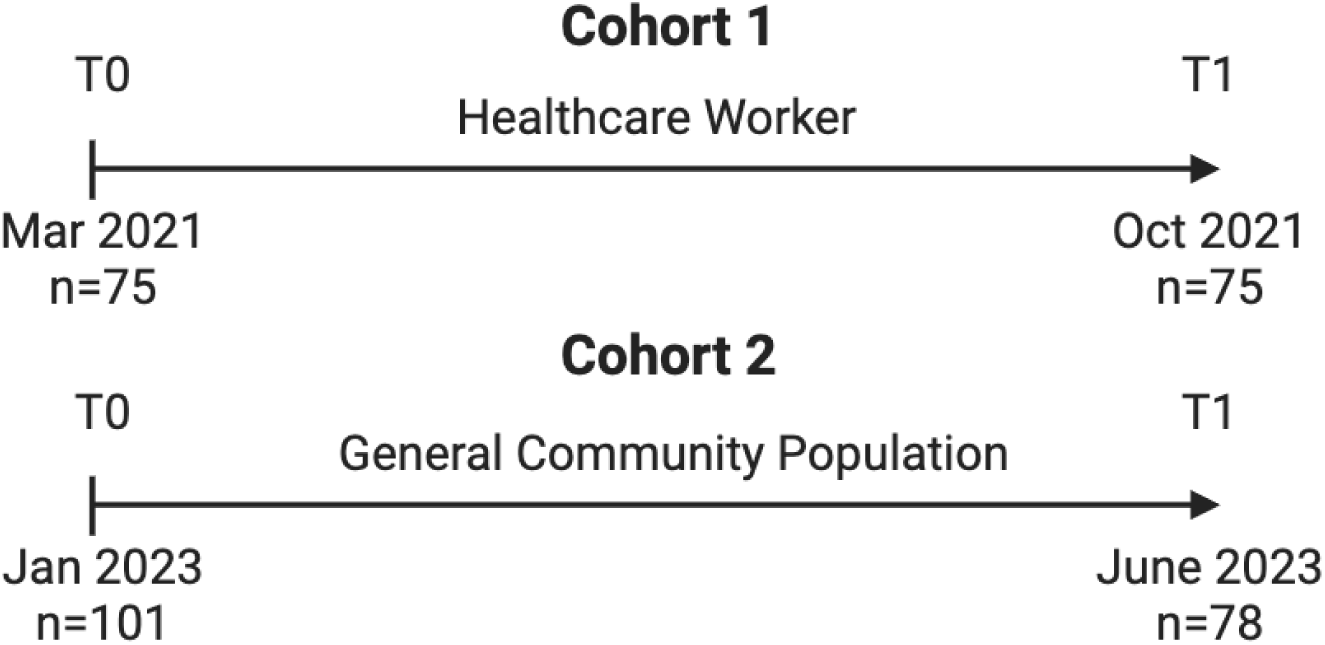
Study design and schematic of study cohorts and patient disposition across two timepoints in **a)** healthcare workers from Lagos, Nigeria and **b)** Abuja, Nigeria from 2023.

**Table 1:** Baseline characteristics of study participants.

|  | All participants | HCW Cohort | Gen Pop Cohort |
| --- | --- | --- | --- |
| **Characteristic** |  |  |  |
| Total number, n (%) | 176 (100) | 75 (43) | 101 (57) |
| Age, median years (IQR) | 37 (29, 43) | 40 (33, 48) | 33 (26, 42) |
| Female gender, n (%) | 94 (53) | 52 (51) | 42 (56) |
| MPXV antibody magnitude-breadth score* | 6140 (3794, 17164) | 6150 (3794, 17164) | 6129 (3473, 13135) |
| Birth cohort**, n (%) |  |  |  |
| Born < 1980 (likely smallpox-vaccinated) | 59 (34) | 35 (47) | 24 (24) |
| Born ≥ 1980 | 117 (66) | 40 (53) | 77 (76) |

\* Magnitude-breadth score derived from the six-antigen targets tested and \*\* Birth-year categories in-line with routine smallpox vaccination which was stopped in 1980

At T0 across both cohorts, using binding antibodies above the antigen-specific cut-off for each target, we observed 48/176 (27%) with no reactivity, 46 (26%) with reactivity to one antigen, 37/176 (21%) with reactivity to two antigens and 21/176 (12%) recognising three antigens (**Table 2; Supplementary Figure 1**). Using a tiered algorithm as previously described^54^ to define our high-confidence seropositivity threshold and to eliminate the likelihood of false positivity from assay artefact or isolated cross-reactivity, we defined MPXV seropositivity as reactivity to at least four of six MPXV antigens. We observed 24/176 (13.6%) showing seropositivity to MPXV comprising mostly of individuals born before 1980 (20/24; 83%) with (4/24; 27%) born after 1980 (**Figure 2; Supplementary Table 1**) and 13/24 (54%) were HCW. We note specifically that only (20/59; 34%) of participants born before 1980 showed MPXV seropositivity suggesting possible low vaccination coverage of ≤34% whilst only (4/101; 4%) of participants born after 1980 showed MPXV seropositivity. Broad responses were uncommon; seven participants (six born prior to 1980) were reactive to five antigens and only three participants (all born prior to 1980) were reactive to all six antigens. We note that most individuals (67/116; 58%) born after 1980 had ≤ 1 reactive antigen (**Table 2**). Additionally, occupational status exerted a lesser effect with 13/75 (17 %) of HCWs relative to 11/101 (11%) community participants meeting the ≥ 4-antigen seropositivity criteria. These baseline patterns highlight both the durability of smallpox-derived immunity in older adults.

**Figure 2.**
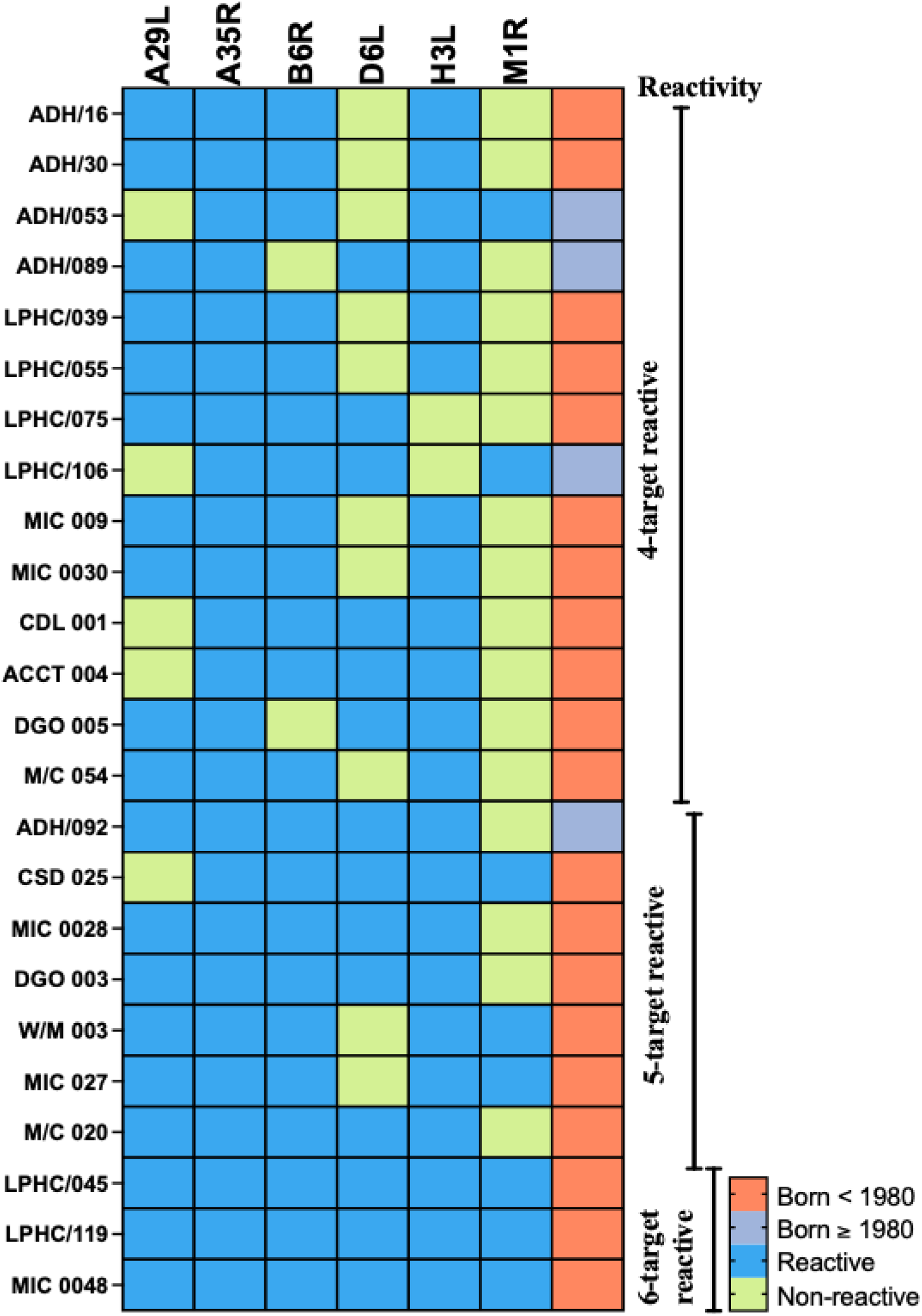
IgG binding profiles of MPXV-seropositive individuals. Binary heat-map showing serum IgG reactivity to six MPXV proteins (A29L, A35R, B6R, D6L, H3L, M1R) in 24 participants who met the criteria for MPXV seropositivity defined as reactivity to ≥4 targets. Each row represents one individual; each column one antigen. Blue squares represent reactive (signal ≥ assay cutoff); light-green squares represent non-reactive participants with seven showing reactivity to ≥5 targets and three showing reactivity to all six targets. The colour-coded side bar indicates individuals born before 1980 (orange squares) or in or after 1980 (grey).

**Table 2:** Distribution of reactive MPXV antigen targets stratified by cohort and birth-year.

|  | Number of reactive antigen targets |  |  |  |  |  |  |
| --- | --- | --- | --- | --- | --- | --- | --- |
| Group (n) | None | 1 | 2 | 3 | 4 | 5 | 6 |
| All participants (176) | 48 (27.3 %) | 46 (26.1 %) | 37 (21.0 %) | 21 (11.9 %) | 14 (8.0 %) | 7 (4.0 %) | 3 (1.7 %) |
| HCW cohort (75) | 22 (29.3 %) | 21 (28.0 %) | 11 (14.7 %) | 8 (10.7 %) | 6 (8.0 %) | 6 (8.0 %) | 1 (1.3 %) |
| Gen Pop cohort (101) | 26 (25.7 %) | 25 (24.8 %) | 26 (25.7 %) | 13 (12.9 %) | 8 (7.9 %) | 1 (1.0 %) | 2 (2.0 %) |
| Born < 1980 (59) | 9 (15.0 %) | 18 (30.0 %) | 5 (8.3 %) | 8 (13.3 %) | 11 (18.3 %) | 6 (10.0 %) | 3 (5.0 %) |
| Born ≥ 1980 (117) | 39 (33.6 %) | 28 (24.1 %) | 32 (27.6 %) | 13 (11.2 %) | 3 (2.6 %) | 1 (0.9 %) | 0 (0 %) |

### Evidence of moderate positive MPXV-specific IgG responses between MPXV-antigens

Following quantification of IgG-specific responses across our study population, we assessed pairwise correlations of log_10_-transformed binding antibody levels for all six antibodies measured (n=176) at T0. Correlation coefficients (ρ) were visualised using a matrix heatmap (**Figure 3a**), alongside pairwise scatter plots illustrating individual-level IgG responses across antigen pairs (**Figure 3b**). We observed moderate and weak positive correlations between multiple antigens, with the strongest correlation observed between the EEV-associated antigens A35R and B6R (ρ=0.65; p<0.0001), consistent withsimilarity. We also observed moderate correlations between A35R and M1R (ρ=0.51; p<0.0001), and between B6R and M1R (ρ=0.53; p<0.0001), suggesting weaker but partially positive pattern between these structurally distinct antigens. All other correlations were relatively weak (ρ range: 0.15-0.43) although statistically significant (p<0.05 for all comparisons). These data indicate a varying but measurable degree of serological correlation across MPXV antigens, with stronger correlations among certain antigen pairs particularly those associated with the EEV form. This pattern of variation supports the potential utility of composite response metrics to capture the breadth of MPXV-specific antibody reactivity which was adapted in subsequent comparative analysis. To ensure this observation was not driven by inclusion of vaccine-naive individuals (i.e., participants born after 1980), we performed a sensitivity analysis restricted to individuals born before 1980 (n=59), i.e. likely to retain residual immunity from smallpox vaccination. The correlation patterns observed remained consistent although a stronger correlation was observed between A35R and B6R (ρ=0.81; p<0.0001) despite some variation in the strength of other pairwise associations. Of note, no significant correlations were observed between D6L and other antigens, indicating that this soluble antigen may elicit a distinct antibody profile.

**Figure 3.**
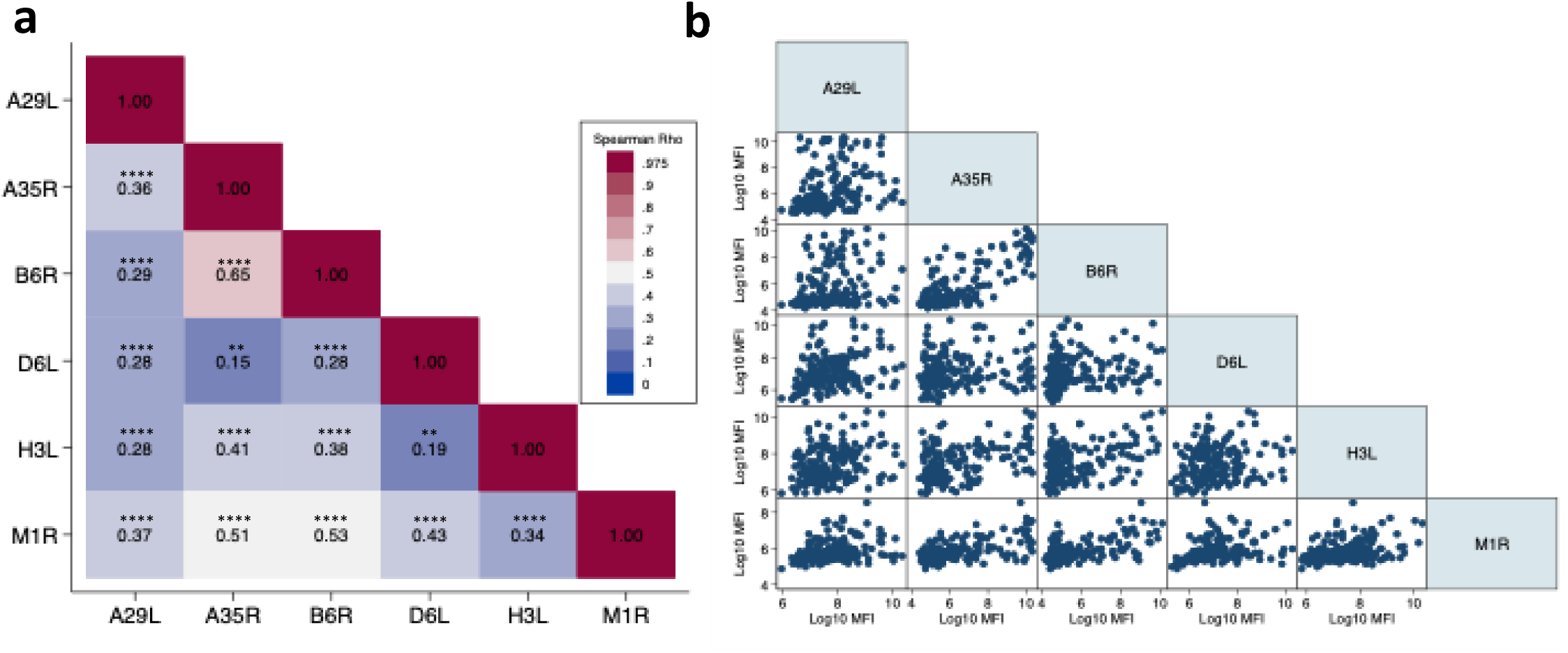
Correlation of MPXV-specific IgG binding antibody responses across six viral antigens. (a) Spearman correlation heatmap matrix of log10-transformed Mean Fluorescence intensity (MFI) values from all HIV negative participants (healthcare workers (n=75) and general population (n=101); total n=176), showing the degree of pairwise correlation (ρ) between antibody responses to six MPXV antigens. The selected targets represent distinct viral structural components: A29L, H3L, and M1R are associated with the intracellular mature virion (IMV); A35R and B6R with the extracellular enveloped virion (EEV); and D6L is a soluble viral protein. The strongest correlation was observed between EEV antigens A35R and B6R (ρ = 0.60) and moderate correlation was also observed between A35R and M1R (ρ = 0.51), and between B6R and M1R (ρ = 0.53) across EEV and IMV targets. Colour shading represents the strength of correlation (ρ), ranging from low (blue) to high (dark red). (b) Pairwise scatter plots of log10-transformed IgG MFI values for each antigen-antigen comparison. Data points reflect individual-level responses. All correlations were statistically significant and are annotated in the heatmap. *P < 0.05; **P < 0.01; ***P < 0.001; ****P < 0.0001.

### Serological profiles suggest Orthopoxvirus residual immunity persists in individuals born prior to 1980

We first sought to assess the role of occupational exposure in shaping MPXV-specific antibody responses. To do this, we measured and compared IgG binding responses across six MPXV antigens in the 176 participants at T0. We observed a relatively higher geometric mean titre (GMT) in responses in HCWs across four of the six antigens tested; A35R (GMT: 726 vs 503), B6R (GMT: 396 vs 255), H3L (GMT: 1904 vs 1522) and M1R (GMT: 401 vs 313), representing ∼1.2-to 1.6-fold higher antibody responses in HCW relative to general population although these differences did not reach statistical significance (**Figure 4a**). In contrast, responses for A29L (2208 vs 2550) and D6L (1129 vs 1318), were marginally lower (∼1.2-fold) in HCWs relative to the general population cohort. These findings suggest that while occupational exposure may modestly influence MPXV seroreactivity, it is unlikely to be a primary contributor to pattern of antibody responses in this setting.

**Figure 4.**
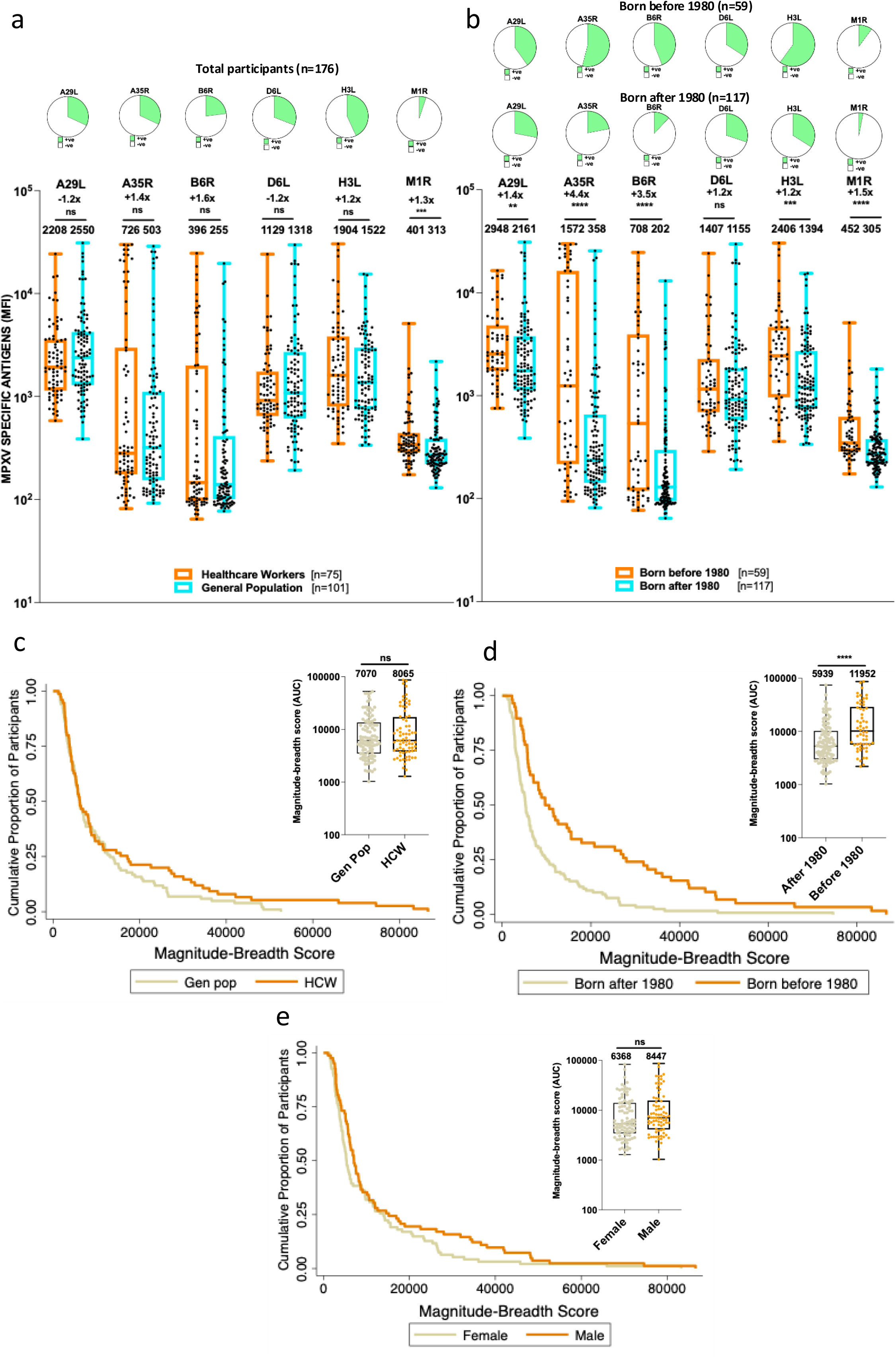
MPXV-specific IgG responses by occupational status and birth cohort. **(a)** Boxplots of IgG binding responses to six MPXV antigens, showing geometric mean titres (GMTs) stratified by occupational status: healthcare workers (HCW, orange) and general population (genpop, blue); and **(b)** by birth cohort: born before 1980 (orange) and born in 1980 or later (blue). The antigens represent distinct viral components: A29L, H3L, and M1R (intracellular mature virion, IMV); A35R and B6R (extracellular enveloped virion, EEV); and D6L (soluble antigen). Boxes indicate the interquartile range (IQR), with horizontal lines denoting medians; whiskers extend to 1.5× IQR. **(c–d)** Cumulative magnitude-breadth plots comparing overall antibody responses across groups. Magnitude-breadth scores represent the sum of binding antibody intensities across all six antigens, capturing both the strength (magnitude) and diversity (breadth) of the humoral response. Panel **(c)** compares distributions by occupational status (HCW vs GenPop); panel **(d)** by birth cohort (born before vs after 1980) and panel **(e)** by gender. Bar charts above panels **a** and **b** shows proportions of participants reactive against each antigen by age cohort. Groups were compared using the Mann–Whitney U test. *P < 0.05; **P < 0.01; ***P < 0.001; ****P < 0.0001.

Next, and given the absence of significant differences in MPXV-specific antibody responses by occupational status, we pooled all the participants (total n=176) stratified by age. We specifically compared individuals born before or after 1980, the year routine smallpox vaccination was discontinued in Nigeria, to evaluate the potential impact of residual smallpox immunity. We observed that participants born before 1980 exhibited significantly higher levels of IgG responses across all six MPXV antigens A29L (GMT: 2948 vs 2161), A35R (GMT: 1572 vs 358), B6R (GMT: 708 vs 202), D6L (GMT: 1407 vs 1155), H3L (GMT: 2406 vs 1394) M1R (GMT: 452 vs 305) in comparison to their younger counterparts (**Figure 4b**) representing ∼1.2-to 4.4-fold higher antibody responses with all targets showing statistically significant difference. These findings are consistent with durable, life-long, cross-reactive antibodies resulting from prior smallpox vaccination. However, the detection of MPXV-reactive IgG antibodies in some individuals born after 1980 suggests possible cryptic exposure to MPXV or cross-reactivity with related Orthopoxviruses, potentially reflecting undiagnosed or undocumented community transmission.

To further validate these findings, we generated magnitude-breadth plots – Kaplan-Meier-style curves reflecting cumulative antibody responses across all six MPXV antigens. These visualisations allow for comparison of the overall magnitude and breadth of antibody responses between groups, where magnitude reflects the strength of binding antibody responses and breadth represents the number of MPXV antigens recognised. Together, this analysis captures both the intensity and diversity of humoral responses against target antigens in each participant. Consistent with the individual antigen analysis is shown in **Figure 4a and 4b**, healthcare workers exhibited a modest rightward shift in magnitude-breadth distribution compared to the general population (**Figure 4c**), though the difference in geometric mean titres (GMTs) of magnitude-breadth score was not statistically significant (score: 14,339 [95% CI: 10,047–18,630] vs. 10,808 [95% CI: 11,376–13,054], p = 0.55). In contrast, participants born before 1980 demonstrated significantly higher cumulative responses than those born after 1980 (**Figure 4d**), with a greater score (19,239 [95% CI: 13996–24482] vs. 8908 [95% CI: 7031–10785], p<0.0001), reinforcing the impact of residual smallpox immunity on shaping MPXV-specific seroreactivity. Notably, we did not find any differences when we compared gender (**Figure 4e**).

### Immunological signatures reveal evidence of recent MPXV exposure

To test the hypothesis of undetected and sustained transmission of MPXV, we employed a longitudinal serological approach leveraging composite measures of antibody magnitude and breadth in the absence of any apparent or reported clinical illness. Using our stringent composite definition, we identified (n=5/153; 3%) participants who demonstrated a ≥2-fold increase into magnitude-breadth score between baseline (T0) and follow-up (T1), in combination with ≥2-fold increases in antibody levels to at least 4 out of the 6 MPXV antigens tested (**Figure 5a, left panel**). These participants showed a significant increase in magnitude breadth between baseline and follow-up GMT (3876 vs 24898; p<0.01) representing a 6.4-fold increase. In contrast, participants who did not meet these criteria only showed a marginal difference (7570 vs 7066; p<0.05) with a negligible fold difference (**Figure 5a, right panel**). These data suggest truly sub-clinical MPXV exposure occurred during the sampling interval in ostensibly healthy individuals which may be captured only through sensitive multiplex antibody profiling rather than routine clinical surveillance.

**Figure 5.**
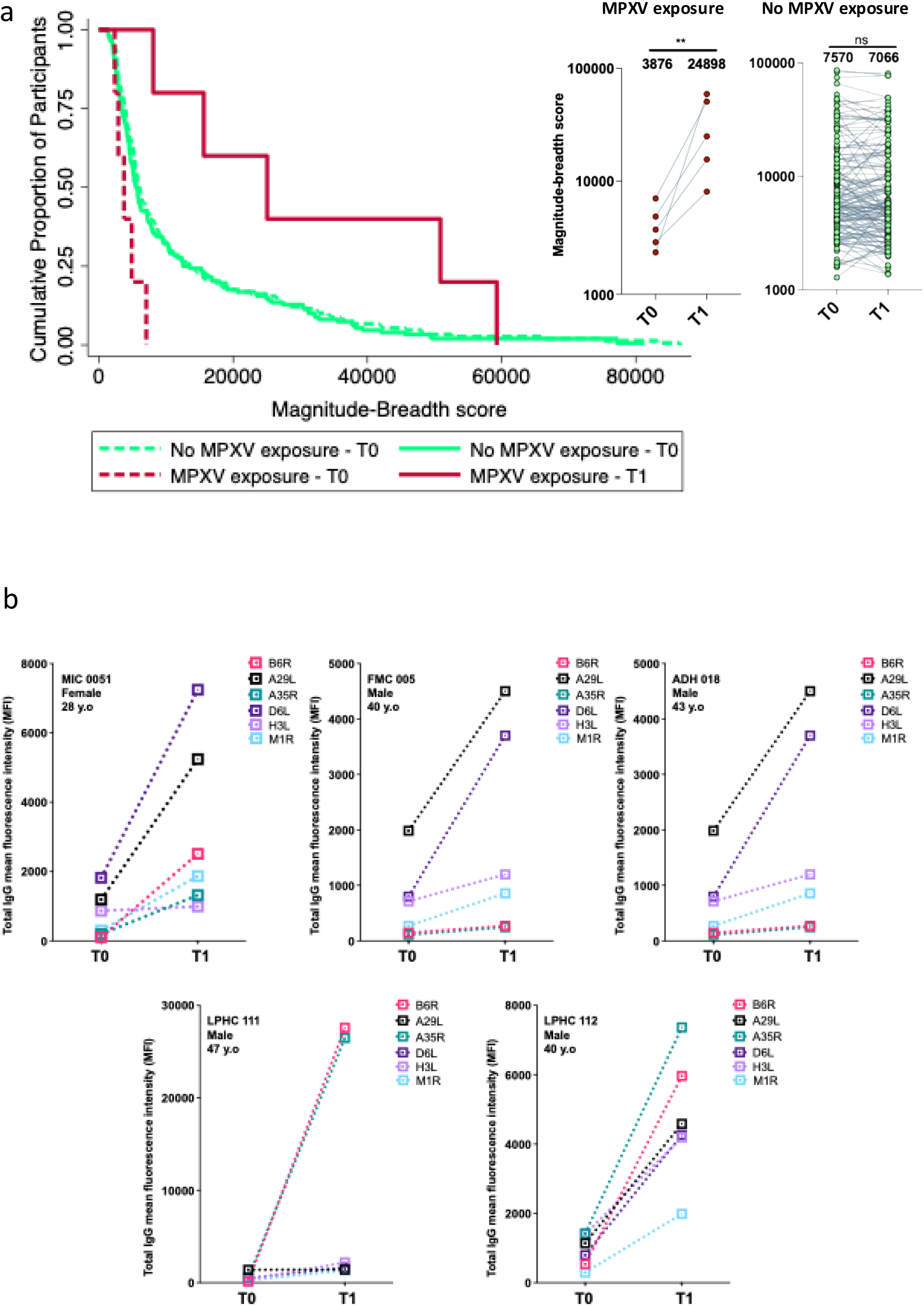
Evidence of MPXV exposure based on composite binding antibody responses. **(a)** Kaplan Meier curves showing magnitude-breadth score comparing participants with and without evidence of MPXV exposure, based on cumulative responses and antigen-specific boosting. **(b)** Paired magnitude-breadth scores from baseline (T0) and follow-up (T1) among individuals classified as exposed and unexposed to MPXV. **(c)** Antibody-specific fold changes in antibody responses across the six MPXV targets (A29L, A35R, B6R, D6L, H3L, M1R) for each exposed participant. Participants were classified as exposed if they demonstrated a >2-fold increase in total AUC between timepoints and ≥ 2-fold increases in ≥4 of the 6 antigens tested.

To further explore potential factors associated with MPXV exposure, we examined individual-level response profiles and antibody kinetics, across the six MPXV antigens (**Figure 5b**). Most participants with MPXV exposure were born before 1980 (3/5; 60%), were mostly males (4/5; 80%) and were all MPXV seronegative at T0. Among MPXV-exposed individuals, the median fold increase in antibody responses between T0 and T1 was highest for B6R (11-folds) and M1R (6.2-fold) followed by A35R (5.2-fold), D6L (4.6-fold), A29L (4-fold) and H3L (2.7-fold). These results indicate that B6R, an EEV-associated antigen, was the most immunodominant target in boosting responses post-exposure. The remaining antigens also showed consistent but varied increases with interquartile ranges (IQRs) of antigenic targets showing substantial individual-level heterogeneity in boosting patterns. This underscores its potential importance in serological surveillance assays for detecting Orthopoxvirus exposure. These data provide additional resolution into the heterogeneity of antibody response and boosting patterns upon exposure/re-exposure to MPXV.

### Genomic supports slow, persistent and covert mpox transmission in Nigeria

To validate patterns of undetected, sustained Mpox transmission indicated by our serological data, we integrated genomic evidence showing a slow epidemic doubling time (∼2 years). ^6,7^ This slow doubling time suggests that viral transmission was limited primarily to specific subsets of the population rather than rapidly spreading across the broader community. To investigate whether a small number of individuals disproportionately contributed to transmission (i.e., superspreading), we measured transmission heterogeneity using the dispersion parameter k, where lower values indicate greater heterogeneity. ^55^ However, it is important to clarify that while transmission heterogeneity alone does not prove cryptic transmission, its presence alongside a slow doubling time implies episodic transmission events occurring beneath the threshold of immediate detection. To quantify Mpox transmission heterogeneity, we analysed the cluster-size distribution of identical sequences collected during the epidemic’s second phase. Given the APOBEC3-mediated mutations of Mpox elevating the mutation rate, we specifically used identical sequences because they indicate very recent transmission events. ^6,7^ However, interpreting identical sequences must be approached cautiously. We acknowledge that identical sequences might represent closely linked transmission chains but may also mask subtle diversification occurring over slightly longer timescales of the genomic data.

We focused on the epidemic’s second phase because this period coincided precisely with the timing of our serological sampling, allowing us to directly integrate genomic and immunological evidence. In total, we analysed 105 sequences ^56^ from this phase (**Figure 6a– b**). The size of these identical sequence clusters ranged from 1 to 3 sequences per cluster, with a median of 7 distinct clusters observed (**Figure 6c–d**). A higher number of smaller clusters suggests many transmission events led to no further spread (i.e., dead-end infections), consistent with substantial transmission heterogeneity. To address ascertainment bias (the likelihood of unreported cases), we conducted sensitivity analyses under two assumptions: first, assuming that half of all reported cases by the NCDC went unreported, yielding a reproductive number (R) estimate of 1.2 (95% CI: 1.1–1.3) and dispersion parameter (k) of approximately 0.4; and second, that all infections were eventually detected (no unreported cases), yielding an r of 1.18 (95% CI: 1.0–1.3) and a similarly low k value of 0.3 (**Figure 6e–f**). Overall, our analyses suggest that the observed transmission heterogeneity alone, irrespective of the reproductive number, likely explains the small polytomy in the second epidemic phase, indicating significant superspreading and frequent singleton dead-end infections This finding aligns with the observed effect of smallpox immunity, acting as an epidemiological bottleneck that constrained broader population-level spread alongside other factors like specific contact-network structures, household transmission all shaped this slow and heterogeneous transmission pattern.

**Figure 6:**
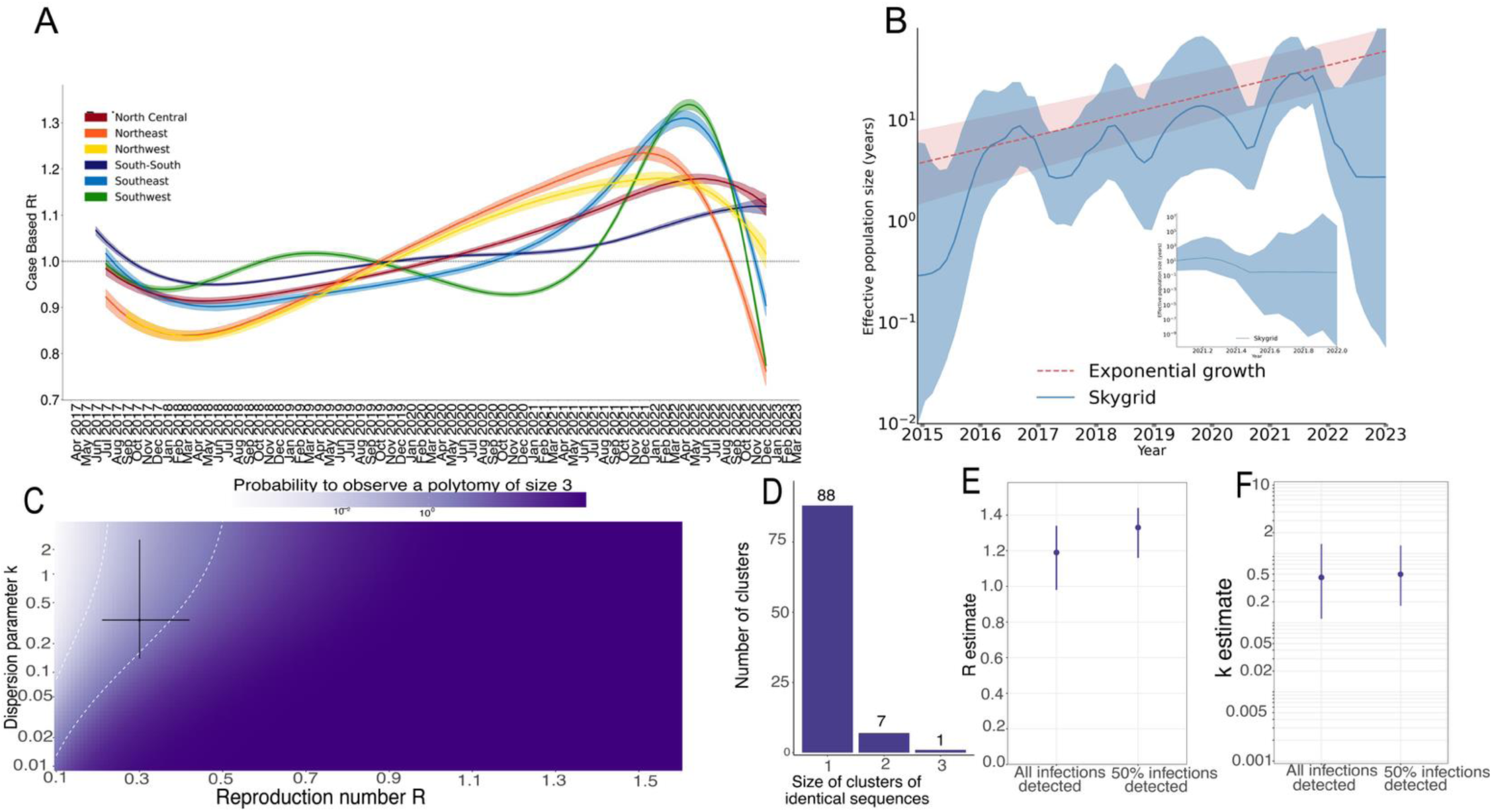
Epidemiological and genomic characterization of Mpox transmission dynamics in Nigeria. (**A**) Estimated reproductive number (Rt) over time, delineating two epidemic phases: initial phase (2017–2018) with Rt < 1, and second phase (2021–2022) with Rt > 1. (**B**) Coalescent reconstruction of the viral effective population size (Nₑ) using a Skygrid model, highlighting transient viral population growth from 2015 to 2017 preceding official case reports, followed by exponential growth during 2021–2022. **C**) Model-derived heatmap illustrating the probability of observing specific cluster sizes based on maximum-likelihood estimates of reproduction number (R≈0.4, 95% CI: 0.3–0.6) and dispersion parameter (k≈0.3, 95% CI: 0.1–0.5). The moderate probability (10–50%) of observing clusters of size three aligns closely with empirical data. (**D-F**) Distribution of cluster sizes from 105 genomic sequences collected during the second epidemic phase, revealing predominantly singleton clusters (average cluster size = 1.1)

## Discussion

Understanding how Orthopoxvirus immunity from historical smallpox vaccination shapes contemporary MPXV susceptibility is critical to estimating population-level risk to infection and guiding rational mpox vaccination strategies, particularly across resource-constrained settings. While smallpox vaccination historically conferred durable, cross-protective immunity to Orthopoxviruses, its global discontinuation post-elimination of smallpox has left a growing global cohort of immunologically naive individuals – particularly those born after WHO eradication vaccination campaigns ended in 1980.^36^ In this context, our findings provide compelling evidence of durable serological immunity in individuals born before 1980, consistent with prior smallpox vaccination^57,58^ and a potentially underestimated yet significant threat^59^: undocumented and silent MPXV exposure without clinical infection in ostensibly healthy individuals suggestive of continuous cryptic transmissions driven by mild or asymptomatic cases that evade syndromic clinical detection.^21^

Our serology data shows residual vaccinia immunity remains broad with a markedly higher IgG magnitude and breadth against MPXV-specific antibodies in individuals born before 1980 relative to their younger counterparts although smallpox vaccination coverage was globally heterogenous with ≍7-60% coverage during the WHO campaigns.^37^ In our cohort, 83% of baseline MPXV seropositive individuals were born before 1980, likely reflective of smallpox vaccination while 60% of younger adults showed ≤1 reactive antigen. These datasets reinforce and extend observations from previous landmark reports characterising the ability of live-attenuated vaccinia virus to induce life-long persisting antibody and T-cell responses persisting for more than 40 years^60–63^ after exposure/vaccination and for up to 80 years.^64^ Our work extends these immunological studies to a West Africa population. Firstly, we demonstrate that residual immunity remains antigen-broad: older participants retained binding activity and reactivity to multiple canonical Orthopoxvirus targets i.e. intracellular mature virion targets (A29L, H3L, M1R) and extracellular enveloped virion targets (A35R, B6R) that structural studies identify as dominant neutralising epitopes.^65^

Next, we observed serological evidence of asymptomatic infection in five ostensibly healthy participants who mounted ≥2-fold increase in magnitude-breadth score and reactivity to ≥ 4-antigen. These individuals were all MPXV seronegative at baseline and mostly males. This observation underscores the protective effect of the historical smallpox vaccination. Our observation of 3% asymptomatic sub-clinical exposure dovetails previous findings from screening cohorts in Spain^12^ with 6% (7/113 participants), France^15^ with 6.5% (13/200 participants), Japan^10^ with 0.4% (5/1346 participants) and Belgium^14^ with 1.3% (3/225 participants). Recent experimental data from the UK provides mechanistic insights into our observations. The data demonstrated that common brown rats (*Rattus norvegicus*) inoculated with MPXV Clade IIb are fully susceptible to infection and upon infection, remain clinically silent despite PCR positivity and continue to shed infectious virus from respiratory tract (within six days) with evidence of serologically conversion^66^ like what we observed. Additionally, the cage-mates of these PCR-positive animals seroconverted without detectable viraemia by PCR.^66^ This study evidence reinforces the possibility of our ≍3% asymptomatic rates at population level and highlights likely cryptic rodent-human/human-rodent/human-human transmission cycles can aid sustain below the radar transmission without clinical symptoms. It is noteworthy that silent low-level transmission is not unique to MPXV. The same cryptic transmission cycle has been observed for arenaviruses such Lassa fever, which is peri domestically maintained by their common African rat (*Mastomys natalensis)* reservoir host. One recent meta-analysis estimated seroprevalence of Lassa fever up to 58% across the West African region^72^ with reported cross-sectional seroprevalence of 50% by age 5^73^ and 82% in a endemic West African population which exceeds reported case numbers; implying substantial asymptomatic spread with limited clinical syndromic capture.^74^ These data indicate higher than reported rates of exposure to heterogenous emerging infectious disease threats that can remain largely undetected.

Our observations are further validated by our country specific phylogenetic reconstruction which revealed how MPXV may persists despite that partial immunity. Cluster-size analysis showed dispersion parameter of k≍0.3, identical to estimates from Europe and the United States during the 2022 Clade IIb epidemic.^75^ Such low dispersion is indicative of frequent dead-end transmission with the outbreaks driven by a minority of superspreaders.^55,56^ Further modelling a 50% under detection, the reproductive number rose only to ∼1.2, and dispersion parameter remained low, implying that even better surveillance would not have fundamentally changed the superspreading-dominated nature of Nigerian mpox transmission. This dynamic aligns with modelling of the 2022 global outbreak and explains how MPXV can circulate at low reported incidence, undetected and yet still generate the boosting serological patterns we observed in our population.^56,75^ In context, the dead-end infection of most mpox transmission means traditional case-based surveillance is unlikely to capture the full extent of mpox transmissions, and public health messaging must be context dependent,^76^ particularly in settings where infections with the Clade IIb presents with mild and atypical symptoms. Furthermore, our focus on serological analysis of samples collected during the COVID-19 when healthcare resources were focused on responding to the pandemic strengthens the plausibility of missed mpox diagnosis.

Antibody-level data in our five participants post-exposure to MPXV highlights the immunodominance of the B6R antigen and its orthologs^77^. This EEV-specific envelope protein expressed in the extracellular virion form of MPXV exhibited the highest post-exposure boost (11-fold) and strongest inter-antigen correlation between antigen pairs. To a lesser extent, M1R (6.2-fold) and A35R (5.2-fold) were also exhibited notable post-exposure boosting. Recent structural and functional studies have highlighted a rationale as to why B6R dominated the boosting hierarchy in participants with recent exposure. Cryo-EM resolution of extracellular B6R core in complex with two B6-specific monoclonal antibodies showed that the B6R antigen is fully exposed on the virus surface with either antibody engaging different sites, suggesting that multiple neutralising sites are simultaneously accessible.^65^ Additionally, both antibodies neutralised extracellular virus in a complement-dependent manner and significantly lowered lung viral load in a lethal vaccinia mouse model suggesting that B6-specific monoclonal antibodies can effectively treat vaccinia infections in this mouse model. ^65^ Further, another mouse model study isolated B6R-specific monoclonal antibody, B7C9, neutralised vaccinia and when administered prophylactically within 24 hours of lethal challenge, significantly reduced lung viral load and weight loss, demonstrating the immunodominance and functional utility of B6R.^78^ It merits attention that a recent study evaluating a mRNA vaccine that encodes a single polypeptide comprising the soluble regions of A35R, M1R and B6R reported 100% protection in mice against lethal vaccinia challenge.^79^ Therefore, these findings align with antigen specific kinetics we observed in participants with MPXV exposure highlighting that B6R in combination with A35R or M1R may be practical surrogate targets for MPXV exposure in unvaccinated populations or be targets for future field-deployable multiplex assays to enhance asymptomatic mpox detection particularly in settings where extensive technical capacity may be limited.

Our analysis was subject to limitations, and we note multiple caveats in our data interpretation. Genomic sampling was geographically skewed toward urban Southern Nigeria, likely under-representing rural transmission and although we assumed 50% underreporting factor in our genomic modelling, actual case detection rates could vary. Our population comprised of COVID-19 vaccinee cohorts where health-seeking behaviour may vary from the general population and potentially increase estimates. Taken together with the above, our analysis measured only binding antibody data rather than functional neutralising antibody data although there is evidence supporting strong correlation between B6R, A35R, M1R and neutralisation. Additionally, serological detection of anti-MPXV IgG cannot completely rule out exposure to other circulating Orthopoxviruses^31^, given antigenic cross-reactivity within the genus^80^ although seropositivity and exposure threshold using our assay required reactivity to ≥4 of 6 MPXV antigens which is relatively conservative but similar to a study which considered reactivity to ≥3 of 5 antigens sufficient and achieved ≈98 % specificity, although waning from vaccination could also have been possible and missed using this cut-off.^54^ Therefore, our higher cut-off therefore maximises specificity and reduces the likelihood of isolated cross-reactivity with other Orthopoxviruses (cowpox, vaccinia-like viruses and camelpox)^31^ which is similar to our earlier work on hepatitis B virus where combining several serological markers minimised false-positivity rates in a high-endemicity African cohort.^80^ Finally, behavioural data was limited which prevented assessment of social drivers of transmission or exposure.

In conclusion, our study suggests that residual smallpox immunity still likely shapes the epidemiological landscape of mpox with older adults born before 1980 retaining broad, multi-antigen reactive IgG profiles with a significant immunity gap relative to individuals born after 1980. The presence of covert, asymptomatic antibody-boosting exposure in a subset of ostensibly healthy participants highlights low-level transmission that symptoms-based surveillance fails to capture.^54,59^ Combining serology-based MPXV-specific targets such as the highly-immunodominant B6R in combination with A35R and M1R surface antigens potentially offers a practical platform for field-deployable tools^54^ to guide precision surveillance and vaccination strategies, particularly in settings where surveillance remains sparse. Future work should i) combine multiplex assay with MPXV and vaccinia neutralisation test to confirm functional protection and ii) prospectively track the dynamics and kinetics of antibody and T-cell immunity following vaccination with new third-generation mpox vaccines such as JYNNEOS to define real-world correlates of immunity in this immunologically diverse population.

## Methods

### Study population and sampling

Our analysis utilised convenience sampling from previous SARS-CoV-2 vaccine studies: *Cohort 1*: Health care workers (HCWs) at the Nigerian Institute of Medical Research (NIMR) and Federal Medical Centre, Ebute Metta, eligible for SARS-CoV-2 vaccination following signed informed consent.^52^ Participants had received their first-dose vaccination between 13 March 2021 and 31 March 2021 and were recruited into the NIMR vaccine effectiveness study. Participants provided plasma samples at baseline (prior to first-dose, T0) and 31months after second dose (T1). Testing was performed on timepoint 0 (T0) and timepoint 1 (T1) samples where available **(Figure 1a**)

*Cohort 2*: HIV Negative general population participants eligible to receive vaccine during the SARS-CoV-2 pandemic at sites affiliated with Institute of Human Virology Nigeria (IHVN) in Abuja.^52^ Eligible participants were men and non-pregnant women >18 years old who had no previous SARS-CoV-2 vaccination and were confirmed HIV negative using the Nigerian national HIV rapid testing algorithm^81,82^. Participants received their first-dose SARS-CoV-2 vaccination between 23^rd^ January 2023 to 20^th^ April 2023. Participants were recruited by i) local community outreach and ii) through phone calls to previously registered patients across three health facilities in Abuja, Nigeria. Testing was performed on T0 and T1 samples where available **(Figure 1b**)

### Laboratory methods and sample testing for MPXV-specific binding antibodies

To capture serological signatures of MPXV exposure given genomic evidence of heterogeneous transmission and potential silent infections, total IgG binding antibody to MPXV-specific antigens were measured in serum samples using a 6-plex the Luminex assay. The selected antigens represent distinct viral structural classes: A29L, H3L and M1R associated with the intracellular mature virion (IMV) form; A35R and B6R associated with the extracellular enveloped virion (EEV); and D6L is a soluble viral protein. For this, six recombinant MPXV proteins were included: A29L (RayBiotech: Cat #230-30237), H3L (RayBiotech; Cat#230-30233), M1R (Sino Biological; Cat #40904-V07H), A35R (RayBiotech; Cat#230-30238), B6R (Sino Biological; Cat # 40902-V08H) and D6R (RayBiotech; Cat#230-01194) were covalently coupled to distinctive carboxylated bead sets (Luminex) to form a 6-plex assay as previously detailed.^83^ The selected antigens represent distinct viral structural classes: A29L, H3L and M1R associated with the intracellular mature virion (IMV) form; A35R and B6R associated with the extracellular enveloped virion (EEV); and D6L is a soluble viral protein. In addition to the antigens, we included BSA (negative-binding control), IL-18BP (mpox encoded virulence factor) and LPS (positive-binding control) and WHO MPXV standards (NIBSC 22/218) as controls. Briefly, beads were activated with 1-ethyl-3-(3-dimethylaminopropyl) carbodiimide hydrochloride (Thermo Fisher Scientific) in the presence of *N*-hydroxysuccinimide (Thermo Fisher Scientific), according to the manufacturer’s instructions, to form amine-reactive intermediates. The activated bead sets were incubated with the corresponding proteins at a concentration of 501μg1ml^−1^ in the reaction mixture for 3 h at room temperature on a rotator. Beads were washed and stored in a blocking buffer (10 mM PBS, 1% BSA, 0.05% NaN_3_). Coupled bead sets were incubated with patient sera at 1/100 dilution for 11h in 96-well filter plates (MultiScreenHTS; Millipore) at room temperature in the dark on a horizontal shaker. Fluids were aspirated with a vacuum manifold and beads were washed three times with 10 mM PBS/0.05% Tween 20. Beads were incubated for 30 min with a PE-labeled anti-human IgG-Fc antibody (Leinco/Biotrend), washed as described above, and resuspended in 100 μl PBS/Tween. They were then analysed on a Luminex analyzer (Luminex/R&D Systems) using Exponent Software V31. Specific binding was reported as Mean Fluorescence Intensity. We defined reactive “cut-off” based on analysis of infant samples (n=14) from Nigeria (presumed MPXV-naive) with mothers born after 1980. Cut-off was defined on antigen-by-antigen basis **(Supplementary Figure 2)**. We defined seropositivity positive cut-off for A29L, A35R, B6R, D6L, H3L and M1R as 3461.5, 814.5, 877.9, 1718.6, 1852 and 1107.2 respectively.

### Magnitude breadth analysis

To characterise cumulative profile of MPXV-specific IgG responses across multiple antigens, we applied a magnitude–breadth analytical framework adapted from prior studies of polyclonal antibody responses in HIV ^84–86^ and SARS-CoV-2 ^87,88^ vaccine studies. This approach estimates the cumulative proportion of participants who retain detectable responses as antigen thresholds increase, effectively capturing both breadth (number of reactive targets) and magnitude (strength of signal) across the tested antigens. Magnitude-breadth curves were constructed using a Kaplan-Meier-like framework, where each participant’s antibody cumulative response to different MPXV antigens was treated as a “time-to-event” outcome with the x-axis representing the magnitude of response (quantified by the area under the curve). Participants with higher magnitude responses across more antigens have a right-shifted curve, reflecting greater immunological breadth. Curves were stratified by participant groups based on occupational exposure, birth cohort and MPXV exposure status. Boxplots of overall magnitude-breadth scores are shown next to the curves. Differences in distributions were assessed using Mann–Whitney U tests.

### Phylogenomic reconstruction of Nigerian mpox spread

To estimate the effective reproduction number (R) of mpox, we adapted the renewal equation framework previously described.^56^ Within this framework, the time varying effective reproduction number, ^89^defined as the average number of secondary infections generated by one infected individual was modelled using a fourth-order spline with five evenly spaced knots. We assumed a discretized gamma distribution for the generation interval, characterised by a mean of 12.6 days and a standard deviation of 5.7 days.^90^ The observed case counts were fitted using a zero-inflated negative binomial distribution to derive posterior estimates of the effective reproduction number as previously described.^91^ To analyse the population dynamics of the mpox epidemic in Nigeria, we adopted an approach similar to that previously described.^6,7^ We utilized recently generated genomic data^7^ within the BEAST software package.^92^ Our analysis employed a non-parametric skygrid coalescent tree prior, incorporating 12 change points distributed over 10 years, as described.^93^ We combined two independent Markov chain Monte Carlo (MCMC) runs, each consisting of 50 million states, with sampling occurring every 2,000 states. The initial 10% of trees from each run were discarded as burn-in. All effective sample size values were confirmed to be above 200.

### Definition of terms and statistical analysis

We compositely defined MPXV exposure as a ≥2-fold increase in total binding antibody magnitude between baseline and follow-up, in combination with a ≥2-fold increase in antibody levels to at least 4/6 MPXV-specific antigens tested. This definition was designed to capture both the magnitude and breadth of the humoral immune response, providing a stringent immunological marker of recent exposure to MPXV or a closely related Orthopoxviruses. Baseline characteristics of participants were summarised as proportions and percentages for categorical variables, and medians with interquartile ranges (IQR) for continuous variables. Antibody responses were visualised using magnitude-breadth curves and paired antigen-specific plots. Distributions of continuous measures between two groups were compared using the Wilcoxon rank-sum or Mann-Whitney test, as appropriate. Reverse cumulative antibody distributions were plotted using Kaplan-Meier survival-style curves, treating area under the curve as the analysis time and exposure group as the failure event.

### Ethics

This study was approved by the Institutional Review Board of NIMR (IRB-21-040 for the Lagos cohort and FCT Health Research Ethics Committee (FCT HREC) for Abuja cohort with approval FHREC/2022/01/193/18-10-22.

## Data Availability

All data produced in the present study are available upon reasonable request to the authors

## Acknowledgement

The authors wish to thank the study participants who made this study possible.

## Competing interests

The authors declare no competing interests.

**Supplementary Table 1:** Baseline characteristics of study participants who were MPXV seropositive at T0.

|  | All participants | HCW Cohort | Gen Pop Cohort |
| --- | --- | --- | --- |
| **Characteristic** |  |  |  |
| Total number, n (%) | 24 (100) | 13 (54) | 11 (46) |
| Age, median years (IQR) | 49 (45, 52) | 52 (46, 55) | 45 (35, 49) |
| Female gender, n (%) | 8 (33) | 5 (38) | 3 (27) |
| MPXV antibody magnitude-breadth score | 31069 (19529, 48158) | 34609 (26750, 65925) | 26260 (18292, 48086) |
| Birth cohort, n (%) |  |  |  |
| Born < 1980 (likely smallpox-vaccinated) | 20 (83) | 13 (100) | 7 (64) |
| Born ≥ 1980 | 4 (17) | 0 (0) | 4 (36) |

