## Supplementary figures for "Legacy immunity from prior smallpox vaccination and serological evidence of asymptomatic mpox transmission in a West African population"

### Slide 1
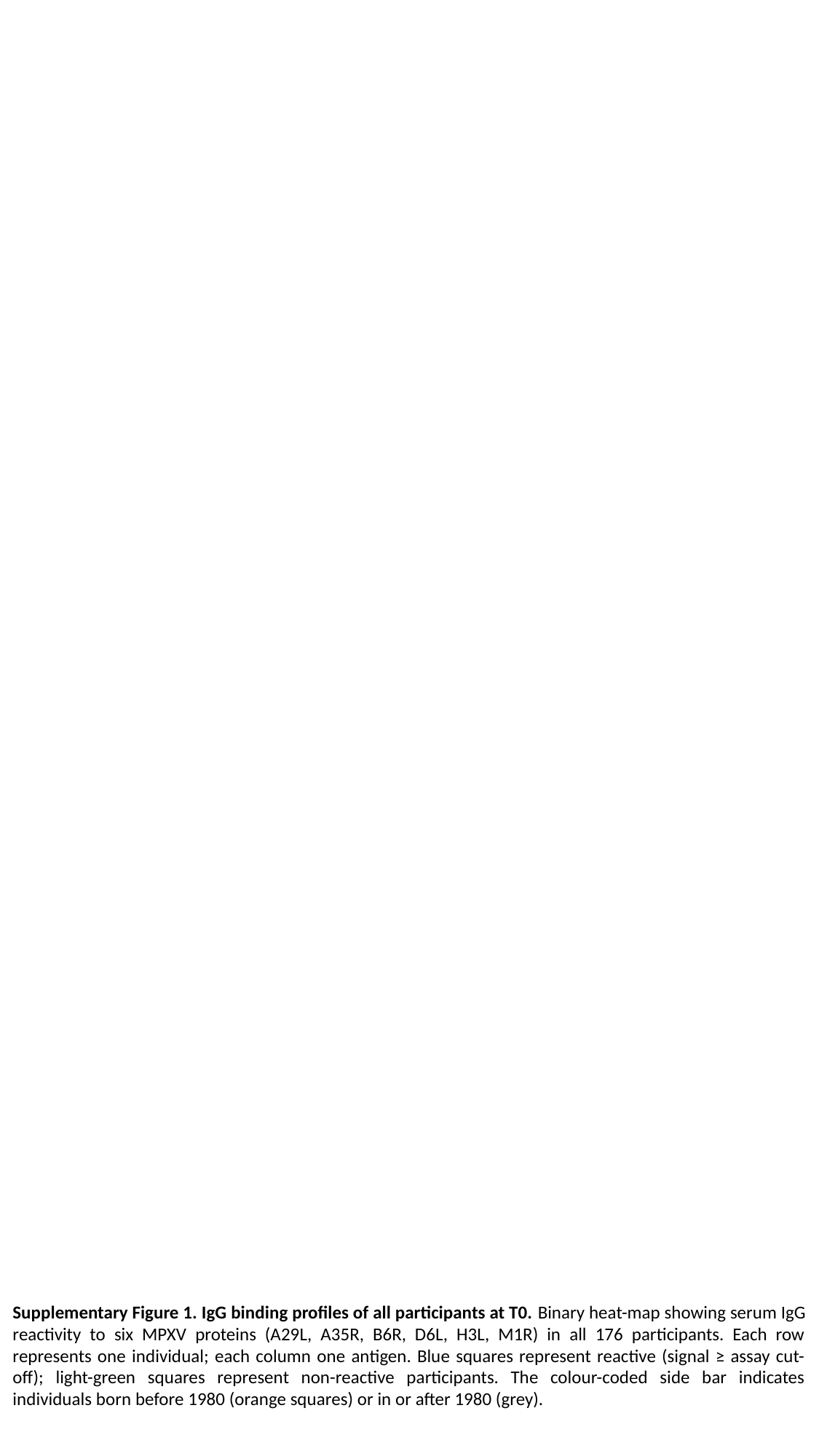

Supplementary Figure 1. IgG binding profiles of all participants at T0. Binary heat-map showing serum IgG reactivity to six MPXV proteins (A29L, A35R, B6R, D6L, H3L, M1R) in all 176 participants. Each row represents one individual; each column one antigen. Blue squares represent reactive (signal ≥ assay cut-off); light-green squares represent non-reactive participants. The colour-coded side bar indicates individuals born before 1980 (orange squares) or in or after 1980 (grey).

### Slide 2
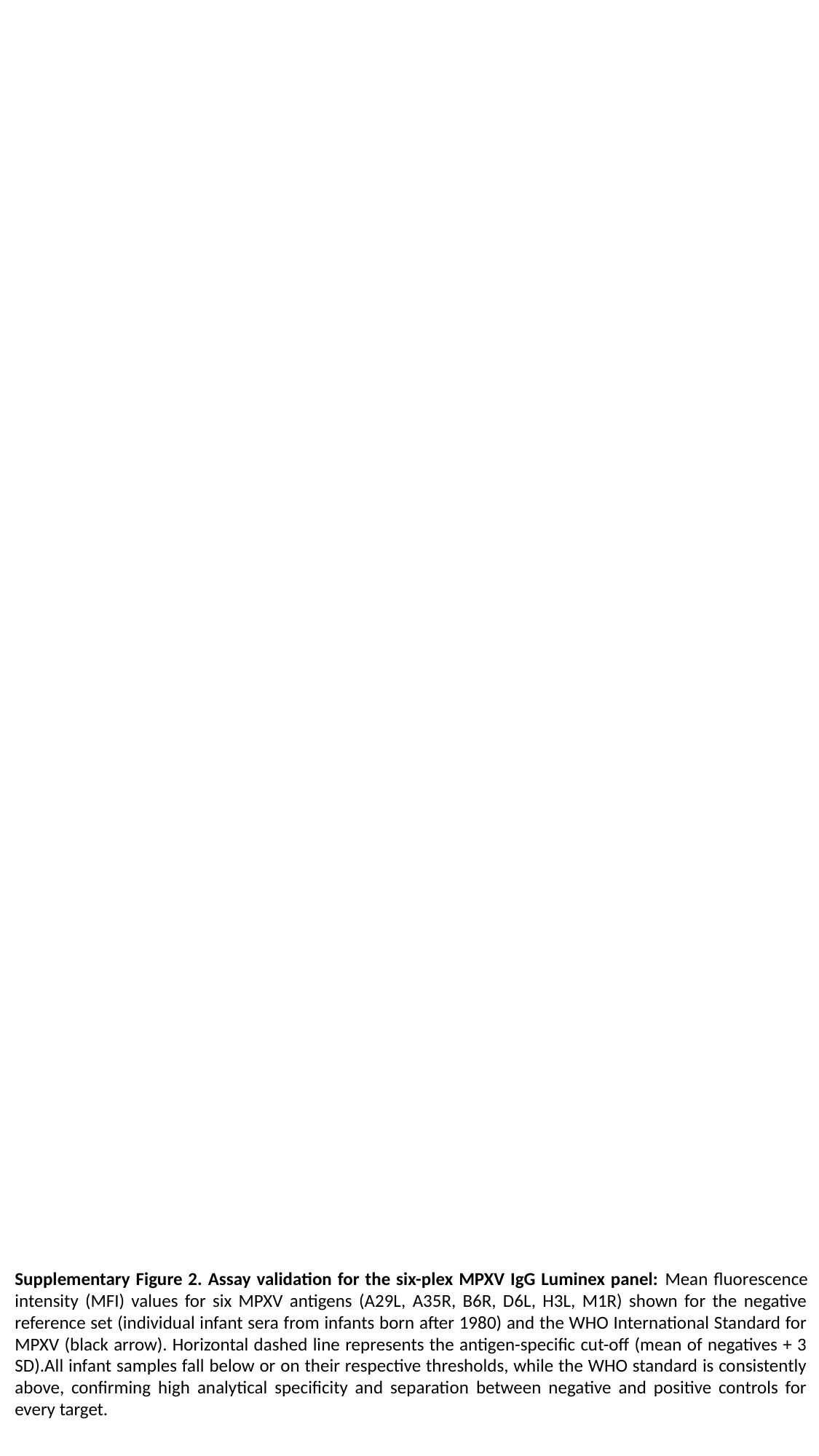

Supplementary Figure 2. Assay validation for the six-plex MPXV IgG Luminex panel: Mean fluorescence intensity (MFI) values for six MPXV antigens (A29L, A35R, B6R, D6L, H3L, M1R) shown for the negative reference set (individual infant sera from infants born after 1980) and the WHO International Standard for MPXV (black arrow). Horizontal dashed line represents the antigen-specific cut-off (mean of negatives + 3 SD).All infant samples fall below or on their respective thresholds, while the WHO standard is consistently above, confirming high analytical specificity and separation between negative and positive controls for every target.
